# Ethnically Diverse Midlife Women’s Menopausal Transition Symptom Experience and Access to Medical and Integrative Health Care: Informing the Development of an Integrative Medicine Group Visit for the Menopausal Transition

**DOI:** 10.1101/2022.06.19.22276301

**Authors:** Lisa Taylor-Swanson, Kari Stoddard, Julie Fritz, Belinda “Beau” Anderson, Melissa Cortez, Lisa Conboy, Xiaoming Sheng, Naomi Flake, Ana Sanchez-Birkhead, Louisa Stark, Nancy Fugate Woods, Paula Gardiner

## Abstract

**Objective:** Individuals in the menopausal transition often seek healthcare in the United States. However, many individuals who seek healthcare do not receive treatments for their symptoms. And, some lack access to providers of both medical care and evidence-based integrative health interventions such as acupuncture, acupressure, or massage. A potential solution to this problem is medical group visits. Medical group visits are when multiple patients are seen by one provider. The present study gathered the opinions of diverse midlife women about interest in and desired design elements of medical group visits for menopause-related symptoms and concerns.

**Methods:** We conducted one focus group with ethnically diverse midlife women to learn about their experiences in the menopausal transition, specifically their symptom experience, barriers, and facilitators to accessing medical and integrative health providers, and their interest in and suggestions for the design of an integrative medical group visit. Qualitative research methods were used to summarize session results.

**Results:** Nine women participated and were diverse in terms of race/ethnicity and religious affiliation, and were highly educated. Themes included: an interest in participating in this conversation; that medical terms were mostly unfamiliar, and that terminology was less important than having a conversation; many symptoms were experienced; social factors affected participants, stressing the need for communication on this topic; receiving both unhelpful and helpful healthcare, a desire for whole person care; a need for information about what conditions Integrative Health interventions can treat, barriers to accessing both conventional and integrative care providers and facilitators include knowledge about insurance coverage and word of mouth. The group expressed great interest in the proposed integrative medical group visit (IMGV) model but expressed barriers such as a lack of time available, and needing childcare. Women indicated that an online format may help to overcome barriers.

**Conclusions:** These findings highlight the importance of engagement with stakeholders before the design and implementation of IMGV and the great need among midlife women for education about the menopausal transition and relevant interventions and self-care.

## BACKGROUND

All individuals with a uterus experience menopause^a^, the cessation of menses, which is a natural occurrence experienced during midlife. In the United States, the mean age of menopause is 51 years^1^. The menopausal transition and early post-menopause is a developmental transition that may last a decade. Even though menopause is a natural occurrence, over 85% of midlife women experience multiple symptoms that interfere with their quality of life and daily activities^2^. Individuals may experience irregular periods, vaginal dryness, hot flashes, chills, night sweats, sleep problems, mood changes, weight gain and slowed metabolism, thinning hair and dry skin, and loss of breast fullness^1,3^.

In addition to the symptoms experienced during peri- and post-menopause, receiving appropriate healthcare can also be difficult. Healthcare barriers affect populations differently. For example, women of color experiencing menopause have a harder time receiving care for their symptoms. One reason for this stems from mistrust in the medical field from generations of maltreatment and lack of treatment^4^. In addition to mistrust in the system, there is a correlation between lower socioeconomic status and higher obesity rates. This connects to menopause because women who are obese are at greater risk of having premenopausal vasomotor symptoms^3^.

The Study of Women Across the Nation (SWAN) was launched because of the disparities experienced by black, indigenous, and people of color (BIPOC) women. The SWAN study is a multi-site, longitudinal, epidemiologic study designed to examine the health of diverse midlife women^5^. The study examines women’s changes during the menopausal transition and early post-menopause concerning physical, biological, psychological, and social changes. This study has informed us that African-American women are more likely to report heavy menstrual bleeding and to undergo hysterectomy^3^.

A common treatment for women experiencing bothersome menopausal symptoms is Menopausal Hormone Therapy (MHT). The Women’s Health Initiative (WHI) conducted a risk/ benefit analysis study of MHT in 2013 and concluded it is appropriate management for some women, while harmful or life-threatening for others^6^. Some of the potential side effects are increased risk for pulmonary embolism, endometrial cancer, coronary heart disease, invasive breast cancer, and stroke. Data were re-examined in 2016 and MHT is listed as a first-line intervention to treat menopausal symptoms such as hot flashes, vaginal dryness, mood changes^1^. Recent studies have indicated minimal to no contraindications MHT utilized with women younger than 60 and during the ten years after menopause^7^. With MHT being a first-line treatment, even though it is not applicable for all women, this creates a care gap as not all women can or choose to take MHT. For example, the State of Menopause Survey conducted with 1039 women ages 40-65 across the United States found that more than 72% of surveyed women were familiar with HRT but 65% said they would not consider using it unless their provider recommended it (32%) or a new clinical study emerged proving its safety (29%).^8^ A systematic review of nine surveys reported that 50.5% of women reported using non-pharmacologic interventions specifically for their menopause symptoms^9^.

Women face many health disparities, high mortality rates, and healthcare access concerns. These problems can partially be attributed to poor education of medical residents^10^. Only 6.8% of surveyed medical residents indicate that they felt adequately prepared to manage the care of women experiencing menopause^10^. Within the medical and nursing educational systems, there is a demand for better resident education regarding care for women in the menopausal transition^11^. Insufficient training translates to poor care: one survey of midlife women reported that one-third of women said they felt their doctor isn’t comfortable talking about menopause, causing them to look elsewhere for support^12^. This could potentially be due to a lack of in-depth education of medical students. Approximately one-third of the surveyed residents opted not to offer MHT to a symptomatic, newly menopausal woman that did not have any contraindications to receiving MHT or to a prematurely menopausal woman until the natural age of menopause, despite the overwhelming evidence that MHT is efficacious and safe for these two categories of women^13^. As previously discussed, MHT does not come without side effects, nevertheless, some medical residents are not providing what is currently the first-line treatment for symptomatic menopausal women. Few publications highlight the attitudes of Nurse Practitioners (NPs) and other healthcare providers toward the management of various symptoms and conditions during the menopausal transition, but literature does exist to educate NPs about the care of women during the menopausal transition^14,15^.

Another barrier is the miscommunication of menopausal terms. Some cultures refer to the perimenopausal term as “the change” or “a transition” instead of using a medical term. In England, menopause is referred to as the “climacteric stage”^16^. In addition, large costs from the healthcare system, lack of insurance coverage, geographic location of residence, life priorities, and native languages are also all barriers that women may face to receiving adequate care.

There are numerous ways to decrease these health disparities. From a clinical standpoint, improving access to health care is a critical solution. Because of the potential negative consequences of MHT and health disparities, more women are turning away from MHT. Group Medical visits are one efficient way to fill this gap.

Group Medical Visits (GMVs), also known as shared medical visits, are medical appointments in which one to two providers deliver medical care for a group of numerous patients with similar diagnoses or questions, instead of a one-to-one appointment. GMVs are one potential solution to the shortage of providers because they allow for more individuals to be seen by a provider trained in concerns of the menopausal transition. GMVs not only allow for individuals to be more easily seen by providers but also creates a community for the patients because they can meet others that are experiencing the same problems^17^. A qualitative study was performed on the delivery of GMVs, concluding that GMVs “successfully deliver on the promise of patient-centered care”^18^.

There is a lack of literature to support that group medical visits (GMVs) are beneficial regarding the topic of menopause and the symptoms women feel. This however does not mean that they are not beneficial for other conditions, or lack the possibility of benefitting midlife women. This topic is currently understudied, as, to the authors’ knowledge, as there is minimal published literature regarding midlife women’s opinions about interest in and acceptability of menopausal transition-focused GMVs. There is much evidence to support the advantages in the care provided at these visits^19^. Of the few studies of GMVs for the menopausal transition, one study conducted on shared medical visits for midlife women included annual examinations, follow-up menopausal concerns, hormone therapy, bone densitometry results, and osteoporosis treatment follow-ups. The study authors concluded that the shared medical visits increased the physician’s productivity by 20% when compared to individual appointments. The participants were surveyed afterward and responded with the majority enduring that they would prefer to go to another shared medical visit instead of the individual visit^20^.

There are promising studies about GMVs for other health topics. A comparison study was conducted with women receiving antenatal care in a GMV setting versus an individual appointment setting and the women preferred group-based antenatal care^21^. For patients who have been diagnosed with diabetes mellitus, the interventions used at shared medical visits improved biophysical outcomes among the patients^22^. Another study asked the question of if psychiatric patients prefer individual outpatient follow-up visits compared to group medical visits. Patients preferred the GMVs to the individual outpatient appointment. While it did not provide a reason for the patient’s choice, it illuminates the desire for GMVs^23^.

The purpose of the present study is to ask ethnically and racially diverse women about their experiences of the menopausal transition, what medical attention they sought out when they had bothersome symptoms, and whether they were interested in GMVs with integrative health components. The results will begin to bridge the gap in knowledge about whether midlife women would prefer GMVs and components of evidenced-based non-pharmacological interventions such as acupuncture.

## METHODS

### Ethics Statement

This study was approved by the University of Utah Institutional Review Board and each participant provided informed consent before the start of the session.

### Community engagement sessions

We held a community engagement focus group^24,25^ (“session”) designed to elicit community members’ opinions about the menopausal transition and access to medical care and evidenced-based non-pharmacological interventions such as acupuncture, acupressure, massage therapy, and chiropractic. We also asked what preferred evidenced-based non-pharmacological components and delivery format might be. Sessions provide a format for researchers to consult with community experts – people who have expertise about a particular topic from their lived experience^25^. The concept of a session is described by the Meharry-Vanderbilt Community-Engaged Research Core^25^ and best practices have been identified to enhance community engagement: early input, researcher coaching, researcher humility, balancing power, neutral facilitator, and preparation of community stakeholders^24^. We incorporated these best practices into the conduct of the studio for which results are presented here.

Our participants were recruited through in-person referrals with the assistance of The University of Utah’s Clinical and Translational Science Institute’s (CTSI) Community Collaboration and Engagement Team (CCET). Recruitment involved flyers, existing community partnerships, word of mouth, social media, and referrals from past participants. The screening was conducted by telephone and email. Potential participants were considered for inclusion if they were female, self-described, other, or prefers not to disclose sex/gender; has an intact uterus; aged 40-55; currently reporting poor menopause-related quality of life (≥3 on a 0-6 scale) and experiencing hot flashes (severity ≥3 on a 0-10 scale) lasting for 6 or more months; willing to provide menstrual history which indicates either late transition (1+ missed periods in the last year) or early post-menopause stage (within 2 years of the final menstrual period); able to provide informed verbal consent.

The following questions were asked in the session:

1. What interested you in this discussion?
2. Are the terms peri-menopause, menopausal transition, and post-menopause familiar?
3. What is your experience with the menopausal transition?
  a. What age is relevant to this topic?
  b. What about social changes or social pressures? Are there any social aspects relevant to your experience of the menopausal transition?
  c. Are there any healthcare aspects to your experience of the menopausal transition?
4. What are barriers to accessing healthcare providers (e.g., primary care providers, gynecologists, etc.)? What are barriers to accessing integrative health providers (e.g., acupuncturists, massage therapists, chiropractors, etc.)?
  a. What helps you access healthcare providers? What helps you access integrative health providers?
  b. Where should resources for the group intervention be posted?
5. What are your feelings about the proposed group intervention?

These questions used lay language and were open-ended when possible. The questions were written to avoid negative or positive bias, and allowed for any amount of detail and self-disclosure. Participants received a short description of the study and the questions at least a week before the session.

The session was held on September 28, 2021. During the 2-hour session, the session coordinator, who was independent of the research team, facilitated a discussion of the question with the aim of eliciting responses from all session participants; a scribe summarized the discussion on large paper as part of the facilitation. Several members of the research team observed the discussion. Sessions were audio-recorded and transcripts derived from the recordings were de-identified. All other data were likewise de-identified. Session participants were provided with a $75 gift card in exchange for their time.

### Data Analysis

Qualitative research methods^26^ were used to summarize the engagement session results. The session was recorded and transcribed. The de-identified transcription was read and re-read, after which thematic coding was performed by the first and second authors, with recoding until consensus was achieved. Participants’ comments are direct quotes, except that they have been edited for brevity while maintaining the original meaning. Descriptive statistics were conducted for the participant demographics using SPSS Version 27^27^.

## RESULTS

Twenty-two participants were screened and nine met inclusion criteria. All nine women chose to attend the session. Participants ranged in ages from 41 to 55 years and the majority had completed college (associates through doctorate degree, 77%) and lived in an urban area (55%). There was broad diversity in terms of race and ethnicity, as well as religion. Other demographic data are listed in Table 1.

**Table 1.**
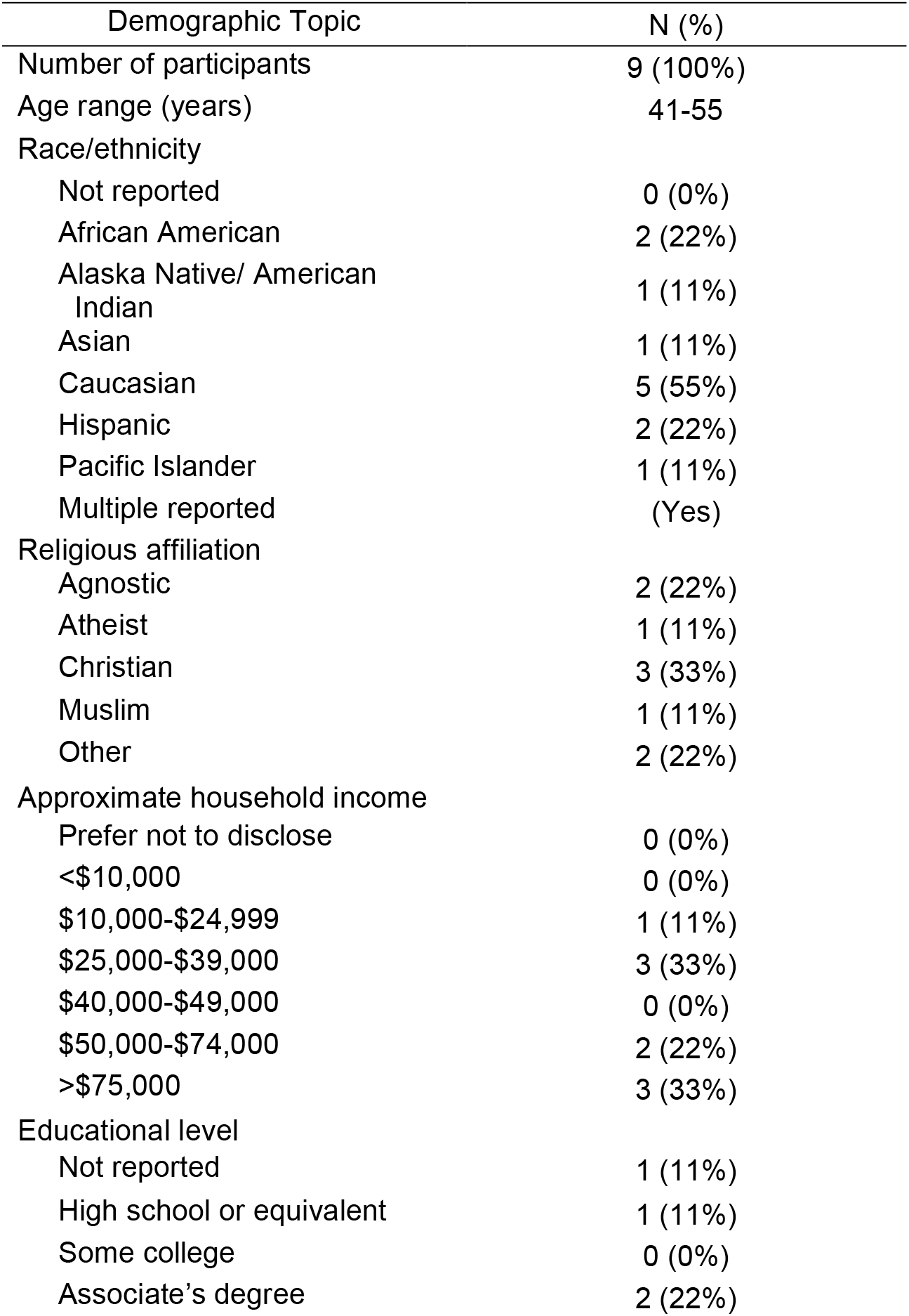
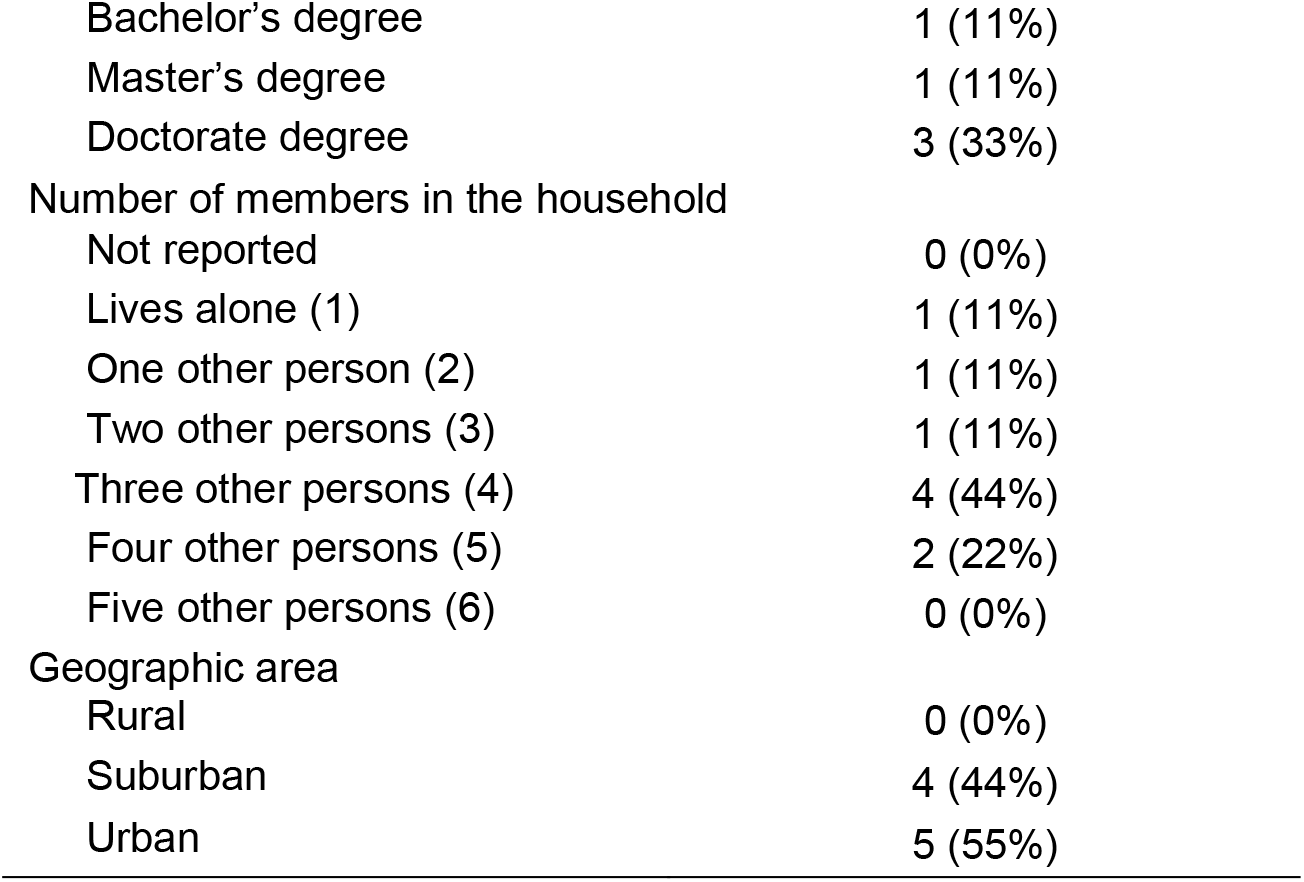
Demographics of engagement session participants

Participants’ responses to the session questions are summarized in Tables 2, 3, 4, 5, and 6. We anticipated that the question “***What interested you in this discussion***?” would elicit participants’ desire to learn more about the menopausal transition, based on the literature^28-31^. This was the case; in addition, participants indicated that they felt alone and would like to talk with other women, in addition to learning more about the menopausal transition (see Table 2).

**Table 2.** What interested you in this discussion?

| Theme | Example Quotes (edited for brevity) |
| --- | --- |
| Interested in this topic | The journey into menopause is very new to me, and I would like to be a part of this important conversation. |
| Learn More | Want to learn ways to get through symptoms and menopause in general.<br>Want to understand what [her] body is doing.<br>Something that she does not know a lot about. |
| Would like to talk with women | Going into menopause and I don't know who to talk to other than a MD.<br>Wanted to be in a space where I could freely talk to other women and share experiences. |
| Interested in integrative health (IH) interventions | Interested in integrative and western medicine and headed into menopause.<br>Heading into the area of menopause and does not usually use traditional medicine health care systems. |
| Not talked about / taboo | My community and society, in general, does not talk about this.<br>Taboo subject in my community and I wanted to step out and learn more and to help out my community. |
| Self-help | Looking forward to this group for self-help options and acupuncture. |
| Feeling alone | Stage in our life's where we feel awkward. Want to hear that I'm not the only one that this is happening to.<br>It's nice to hear from women who are also experiencing the same thing. |

**Table 3.** Are the terms peri-menopause and post-menopause familiar?

| Term Used | Example Quotes (edited for brevity) | Culture (if stated) |
| --- | --- | --- |
| Peri-menopause | I had never heard of perimenopause until joining this group. | -- |
|  | I feel that it encompasses the pre, actual, and post. |  |
|  | I'm in health research and I've never come across that term [perimenopause] before. | -- |
| Post-menopause | Post menopause: I am not familiar with that term at all. | -- |
| Which terms to use | The terms are not as important as the conversation. | -- |
|  | One is not better, they're all important. | Latino |
| "The change of life" | Almost like a timeline. Although it isn't really talked about so I'm still trying to figure out what age is the timeline. | Polynesian |
| "Transition" or<br>"Transitioning" | I like it because I am becoming something else, although there are a million different ways we transition and one term does not acknowledge this uniqueness. | -- |
| "The change" | Feels like a colloquial term. | -- |
| No word for it because<br>taboo subject | Familiar as in hearing about them, but what does it mean? | Somalia |

**Table 4.1.**
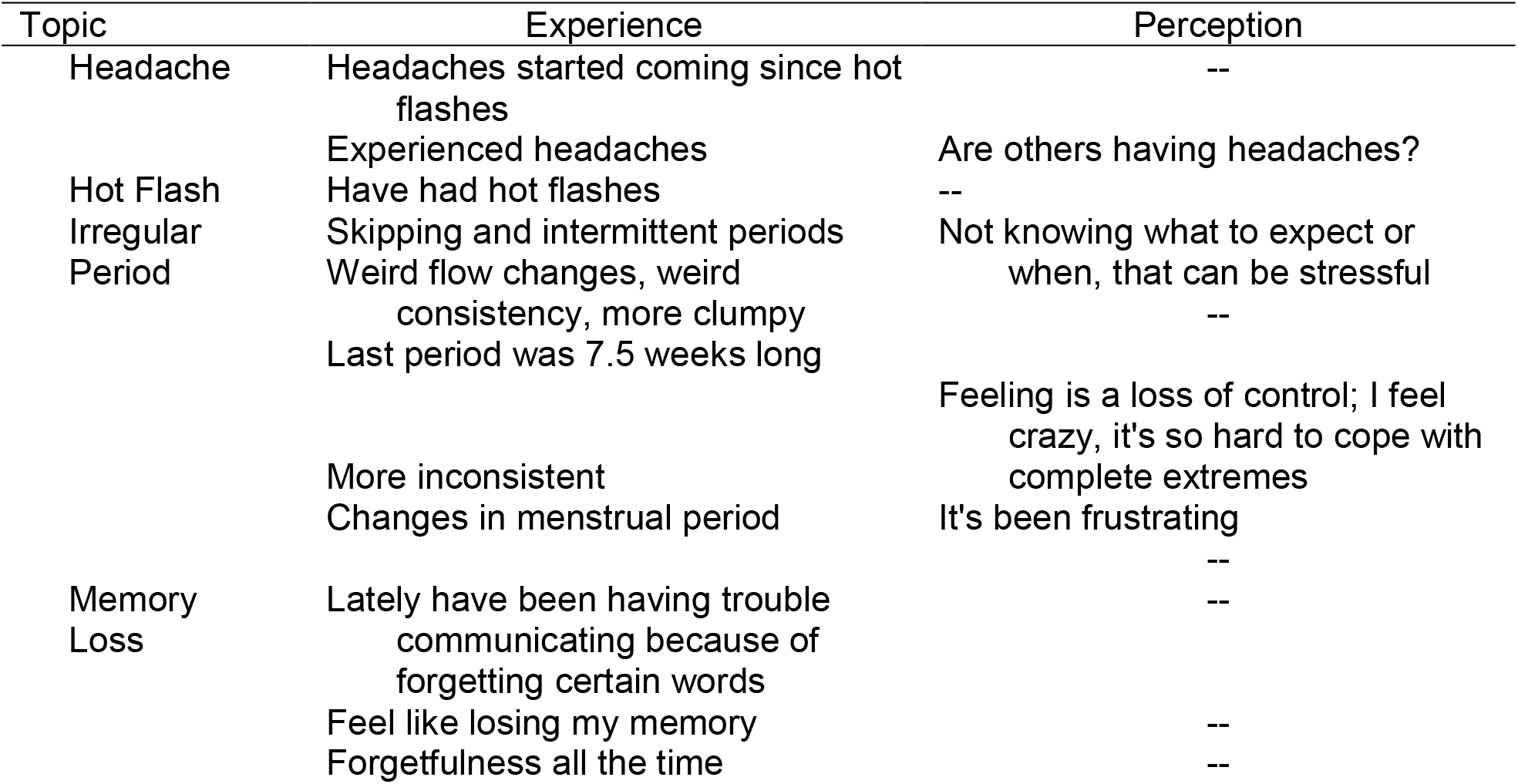
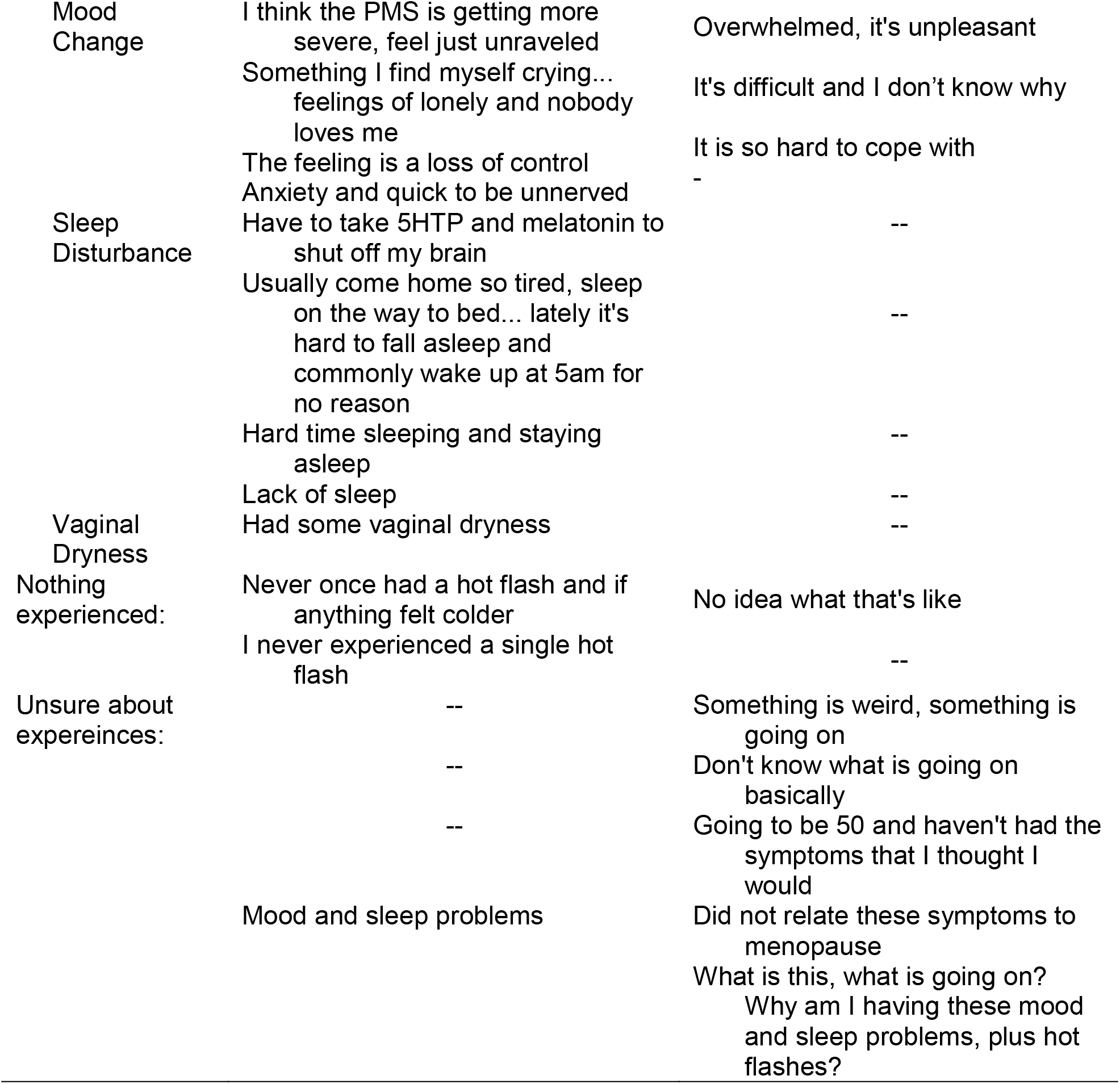
What is your experience with the menopausal transition?

**Table 4.2.** Health Care Dimensions

| Age | Onset of Symptoms |
| --- | --- |
| 39 | Intrauterine device removed, and periods have been irregular since. |
| Early 40's | Skipping and intermittent periods |
| 41 | Having troubles with mood and sleep |
| 42 | Never experienced a hot flash, so maybe it's on its way? |
| Mid 40's | Period stopped |
| 50 | I'm going to be 50; I haven't had the symptoms I thought I would. |

**Table 4.3.** Social Dimensions

| Occurrences | Interactions, Thoughts, and Feelings |
| --- | --- |
| Participants in the "Sandwich Generation" | Taking care of elderly parents and kids in middle school |
| Participants trying to confide in their friends | They have no idea what I am talking about.<br>Friends do not know anything about menopause. They make fun of her and may say things like “oh you're old”<br>I currently do not have friends who are experiencing any symptoms, do not have friends who are experiencing any symptoms, do not really have anyone to talk to about this. |
| Anticipating menopausal experiences from mothers | Mom had a hysterectomy in her 40s, did not have perimenopause either. Hers was immediate. |
| Communication about the menopausal transition | Communication is needed in the community and between community people and their providers. |

**Table 4.4.** Age Dimensions

| Themes | Participants' Perceptions of the Listed Topics |
| --- | --- |
| Unhelpful care from a provider | The doctor never associated their symptoms with menopause.<br>The doctor thinks their symptoms have something to do with medication.<br>Frustrating with listening to her doctor when she goes in. Seems like 15-minute office visit with a doctor because of symptoms, then blood work, and the doctor said that they are fine and instead they feel "crazy." The doctor has not associated symptoms with menopause or had a conversation with her and she is 50 years old. |
| Helpful Care from Provider | The Nurse Practitioner was profoundly different than with male gynecologist, it was helpful, and she treated her as a whole person.<br>Gynecologist she saw was great, but only had "medical fixes."<br>Participant appreciated that her male health care provider immediately asked if she was having irregular periods regarding her symptoms of trouble sleeping and mood changes. |
| Sex of the provider | More comfortable talking to women providers that are within the menopausal range versus that of being in their 20's or being male.<br>A male doctor ran bloodwork and said the participant was not in perimenopause even though she had many symptoms. |
| Pharmacologic fixes versus whole person care | Am I in need of a change of dose? Change medication?<br>There should have been a discussion before giving a different medication.<br>She just thought she needed a change of medication.<br>Not just about the medication or prescription; it is about helping a whole woman through this process. |

**Table 5.**
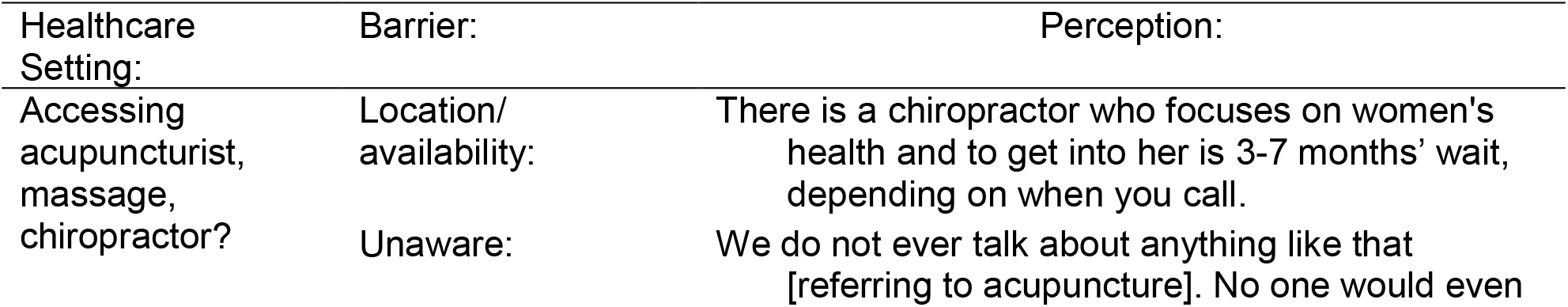
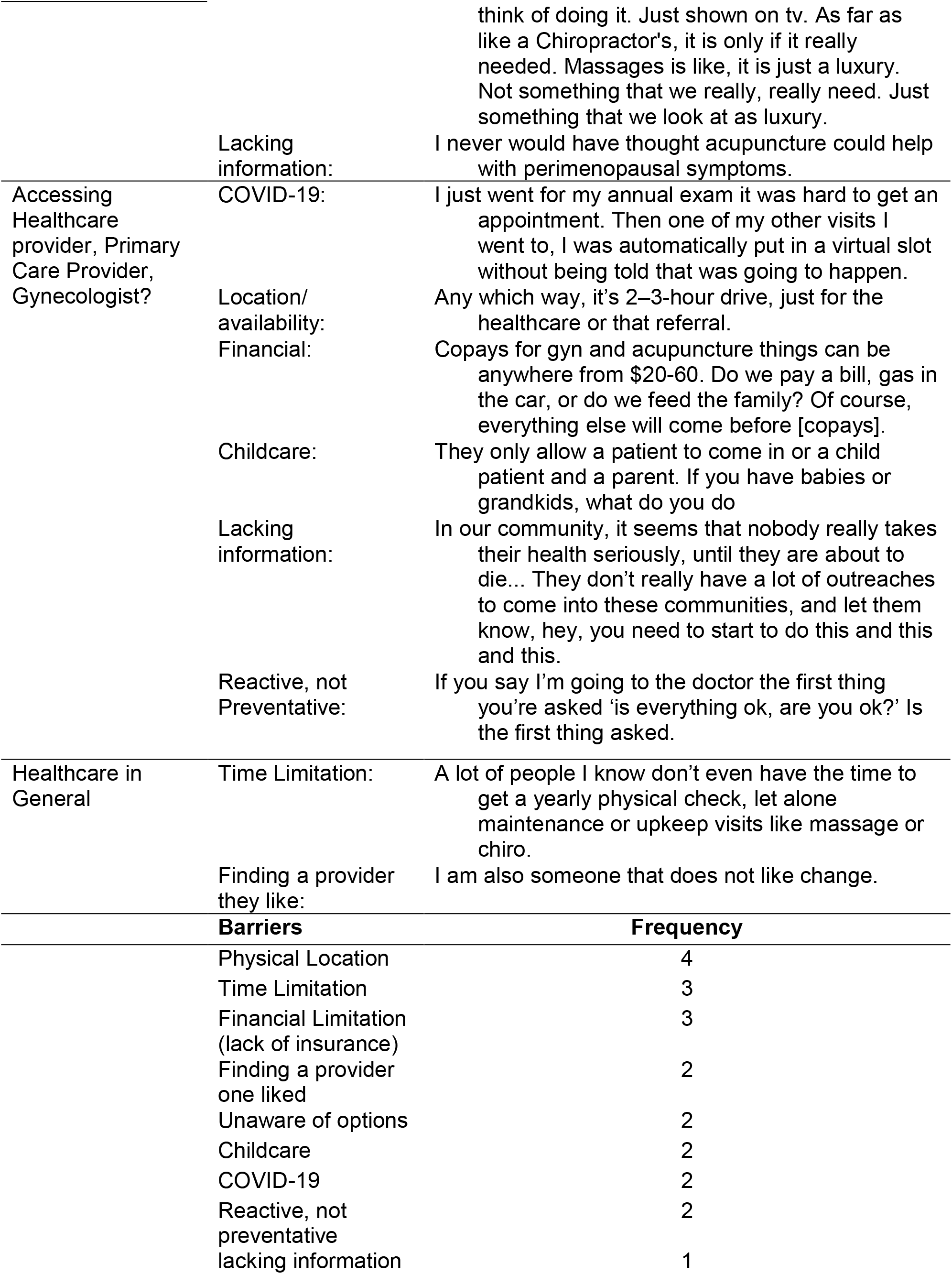
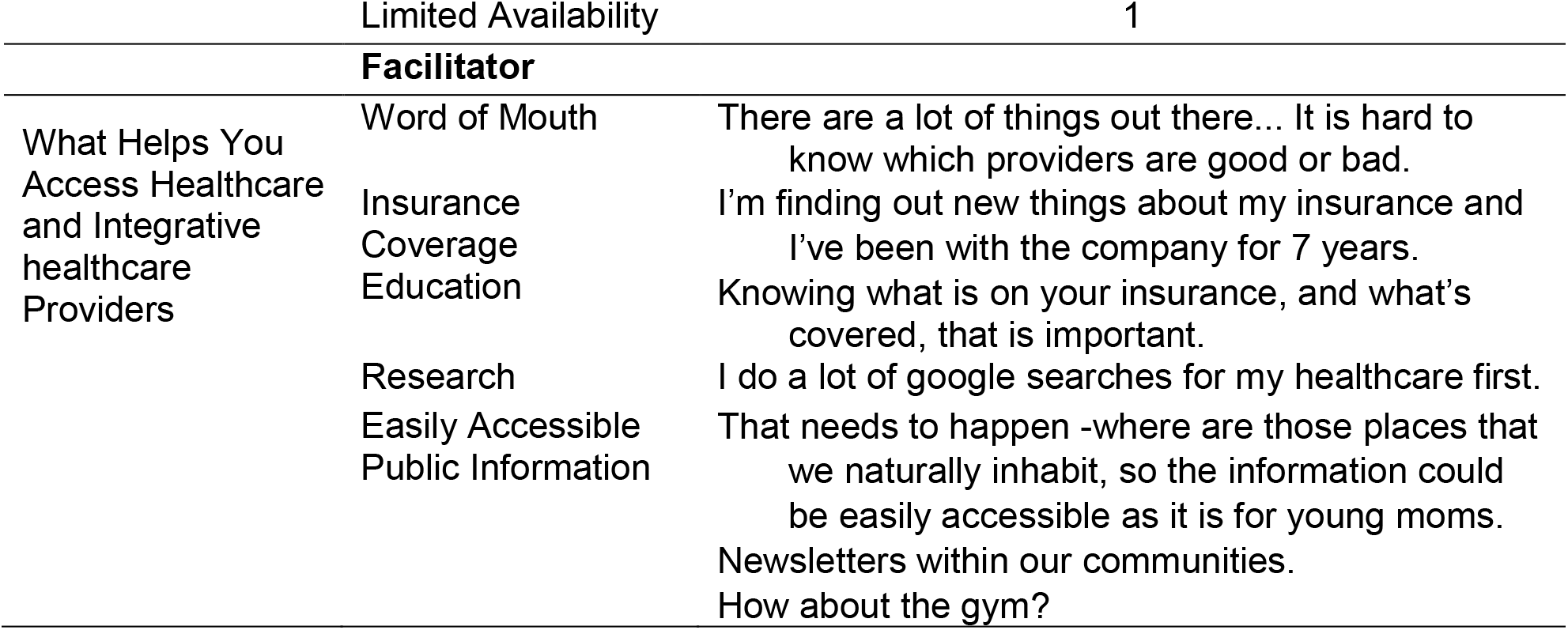
Barriers and Facilitators to Accessing Healthcare

**Table 6.** Perceptions of the Proposed IMGV Model

| Themes | Input about IMGV |
| --- | --- |
| Positives | We have this designated area to talk and its safe and nice to hear that others have similar or different experiences. Having a designed area and time to set aside to do this seems like a fantastic idea. |
|  | I like that there is actual maintenance after. A lot of time through a program and often on your own. Like that aspect.<br>When you started talking all I could think was can I sign up today?! That is the most profound thing.<br>I like that is a whole person approach that addresses a wide variety of needs. But most importantly, that women are actually being seen. That is the key, in a lot of spaces, we are not feeling seen. When almost all the women go through this in their lives. So a huge part of the population never feels seen and addressed.<br>The most valuable thing that this program seems to be addressing, is that acknowledgment. The value of having space for a discussion.<br>It's in-depth teaching and amazing. Grateful that it is going to be something available for a lot of us.<br>I am definitely getting lots of different ideas that could be possibilities that I could integrate in our community. Just in the couple of hours that I have experienced tonight. |
| Barriers/<br>Additions | Definitely lack of free time is a barrier and it's an investment we will need to figure out – but I will do whatever I can to be there.<br>Many women have small children. Maybe add childcare somehow. Then get a lot of women interested in having 2-3 hours of space without having to worry. If barriers were removed, I think women absolutely could participate and would want to.<br>I just wish if you can also maybe put in cultural and language barriers. Maybe add Different language, reach out to different cultures. That would be wonderful.<br>I feel a little bit overwhelmed by it. ... To find time, journaling, and trying to eat healthy and all of that. Just to me the whole thing I kept thinking, 'would I even have time to do all that??' I would love to join it but I don't know if I could really commit to it. |
| Format for IMGV | I love online. If I have to travel, bad weather, construction, traffic... I like zoom.<br>Maybe a/synchronous, alternating may be beneficial.<br>CANVAS<br>Discussion boards: done on your own time, in your own space. Not live. |

### Are the terms peri-menopause, menopausal transition, and post-menopause familiar?

Several views were presented concerning terminologies such as perimenopause, menopausal transition, and postmenopause (please refer to Table 3). Several women reported that they had not heard of various terms before attending this session. Women also noted that the terminology is less important than having conversations on the topic; other women noted terms such as ‘change of life’ or ‘transition’ were helpful.

Tables 4.1, 4.2, 4.3, and 4.4 include the following topics: women’s experiences with the menopausal transition; ages relevant to this topic; social dimensions; healthcare dimensions.

### What is your experience with the menopausal transition?

With respect to experiences, participants reported experiencing headaches, hot flashes, irregular menstrual periods, loss of memory, changes in mood, sleep disturbances, vaginal dryness, as well as no symptoms being experienced, and being unsure about experiences. See Table 4.1 which also includes perceptions, or appraisals, about the various symptom experiences.

### What age is relevant to this topic?

There were also age dimensions mentioned by participants. Some women reported that they had irregular periods in their late 30’s and 40s, and had problems with mood and sleep, as well as some women reported that they did not have particular symptoms in their 40s and did not have the symptoms they thought they would in their 50s (see Table 4.2).

### Are there any social aspects to your experience of the menopausal transition?

Social dimensions reported by women (see Table 4.3) included being in the “sandwich generation”, trying to confide in friends but finding they were not also in the menopausal transition, basing anticipated menopausal experiences on their mother’s experiences, and that communication was needed about the transition.

### Are there any healthcare aspects to your experience of the menopausal transition?

Participants reported healthcare dimensions (see Table 4.4) such as unhelpful care from a provider, helpful care from a provider, the sex of the provider being relevant, and interest in whole person care versus pharmacologic fixes.

### What are barriers to accessing healthcare providers (e.g., primary care providers, gynecologists, etc.)? What are barriers to accessing integrative health providers (e.g., acupuncturists, massage therapists, chiropractors, etc.)? What helps you access healthcare providers? What helps you access integrative health providers? Where should resources for the group intervention be posted?

Participants provided detailed comments regarding barriers and facilitators to accessing health care – both conventional healthcare providers and integrative healthcare providers, such as acupuncturists, chiropractors, and massage therapists (see Table 5). Concerning accessing Integrative Health care providers, participants noted the location and availability were an issue, as well as a lack of awareness about what acupuncture or massage could be used to treat. With respect to accessing conventional care, COVID-19 related concerns were noted, as well as issues with location and availability of the provider, need for childcare, lacking information about going in for preventive care, but also finding that providers are reactive and not preventative. General limitations were noted such as time and finding a provider they liked. A frequency count of barriers is included in Table 5, with the physical location being the most frequently mentioned barrier.

Facilitators to accessing healthcare providers (both conventional and integrative) included word of mouth, having insurance coverage, education about the insurance plan, doing their research, and having easily accessible public information (along with suggestions as to where we could post information about the proposed IMGV).

### What are your feelings about the proposed group medical visit intervention?

Participants’ perceptions of the proposed integrative medical group visit (IMGV) are listed in Table 6. Participants indicated several *positives* about IMGV, such as a safe and designated time and place to talk about the menopausal transition, desire for a follow-up session, excitement about the IMGV, and that it is a whole person approach. Another expressed theme was that women feel unheard and unseen at this age and in this transition and that the IMGV would give acknowledgment to women. Several *barriers* were described, such as midlife women being very busy and lacking time to devote to a multi-session intervention, that childcare may be needed, and that perhaps the IMGV would need to be tailored for various cultures (and delivered in various languages), and that the IMGV might be overwhelming with many different components such as journaling and eating healthily. Suggested formats included online, either synchronously or asynchronously with a learning management system such as Canvas or via discussion boards.

We conducted a Return of Results on May 19, 2022, and 6 of 9 participants attended. The research team reviewed the data tables presented here with midlife women. Participants provided input regarding clarifications and confirmed the identified themes in each of the tables. Feedback on table 4 included that after the discussion in the fall, women’s thinking changed, and that hearing other women’s perspectives changed their own perspective. Several women mentioned that they had sought care from a healthcare provider; one participant found a menopause group on Facebook and found the discussion there helpful. Participants mentioned that having community-specific tailoring to the IMGV would be appropriate and that the biggest identified need is education for midlife women about this topic because every woman will someday go through menopause.

## DISCUSSION

The community engagement session with nine midlife women yielded important information about their lived experiences during the menopausal transition. Participants were diverse in terms of race/ethnicity and religious affiliation and were highly educated. Themes included an interest in participating in this conversation; that medical terms of peri-, post-menopausal and menopausal transition was mostly unfamiliar, and that terminology was less important than having a conversation on this developmental transition; many symptoms were experienced by this sample of midlife women; that ages 39-50 are relevant to this topic; many social factors affected participants, stressing the need for communication on this topic; receiving unhelpful and helpful healthcare, a desire for whole person care; a need for information about what conditions Integrative Health interventions can treat, barriers to accessing both conventional and integrative care providers and facilitators include knowledge about insurance coverage and word of mouth. The group expressed great interest in the proposed integrative medicial group visit (IMGV) model but expressed barriers such as a lack of time available, and needing childcare. Women indicated that an online format may be helpful to overcome these barriers. These findings highlight the importance of extensive engagement with potential stakeholders before the design and implementation of IMGV are conducted.

Our findings are in alignment with existing literature on women’s experiences of the menopausal transition. Unfortunately, the same concerns expressed by a focus group conducted over a decade ago were echoed in the session we conducted: lack of support and confusion about symptoms attributable to the menopausal transition, as well as difficulties in obtaining helpful care from healthcare providers^31^. Midlife women want reliable information and opportunities to discuss the menopausal transition with (preferably female) health professionals is another theme from the present study that aligns with prior literature^32^.

Although the session provides important preliminary information about midlife women’s beliefs about peri-menopause, the menopausal transition and post-menopause, symptoms experienced, and access and barriers to medical and integrative care, there are limitations associated with our study. We had a slightly younger sample, and we did not have any participants who post-menopausal. We also had a highly educated sample living in urban and suburban, thus results may not be representative of women living in rural or frontier areas, as well as those with less education. Study strengths include a sample with diversity with respect to race, ethnicity, and religious affiliation.

Our study suggests the great need for education by midlife women about the menopausal transition – information should include symptoms commonly experienced, the timing of symptoms during the stages of the transition (late reproductive, early and late transition, early and late postmenopause), those symptoms may last for years for some women while others are asymptomatic or experience minimally bothersome symptoms. Further, education should include self-care symptom management tips, pharmacological and non-pharmacological care, and information that is culturally relevant and tailored. Our participants repeatedly stressed that the social nature of the session was helpful and that women felt reassured they “weren’t going crazy” because other women in the group expressed having similar experiences. There is a great need for information and support for midlife women in the menopausal transition. The social support, education, and self-care included in an IMGV may provide much-needed care during this natural, but sometimes problematic, life transition experienced by everyone with a uterus.

## Data Availability

All data produced in the present study are available upon reasonable request to the authors

## Acknowledgments

The research team would like to thank the participants for their time, expertise, and input.

## Footnotes

a While menopause is typically discussed as a cisgender women’s health topic, not all people identify as cisgender women who experience menopause. Therefore, in this article we use the terms “individuals experiencing menopause” and “experiences” instead of “symptoms” to avoid pathologizing/medicalizing menopause.

